# Cross-Field Strength and Multi-Vendor Validation of MagDensity for MRI-based Quantitative Breast Density Analysis

**DOI:** 10.1101/2024.12.08.24318677

**Authors:** Jia Ying, Renee Cattell, Chuan Huang

## Abstract

**Purpose:** Breast density (BD) is a significant risk factor for breast cancer, yet current assessment methods lack automation, quantification, and cross-platform consistency. This study aims to evaluate MagDensity, a novel magnetic resonance imaging (MRI)-based quantitative BD measure, for its validity and reliability across different imaging platforms.

**Methods:** Ten healthy volunteers participated in this prospective study, undergoing fat-water MRI scans on three scanners: 3T Siemens Prisma, 3T Siemens Biograph mMR, and 1.5T GE Signa. Great effort was made to schedule all scans within a narrow three-hour window on the same day to minimize any potential intraday variations, highlighting the logistical challenges involved. BD was assessed using the MagDensity technique, which included combining magnitude and phase images, applying a fat-water separation technique, employing an automated whole-breast segmentation algorithm, and quantifying the volumetric water fraction. The agreement between measures was analyzed using mean differences, two-tailed t-tests, Pearson’s correlation coefficients, and Bland-Altman plots.

**Results:** No statistically significant differences in BD measurements by MagDensity within the same field strength and vendor (3T Siemens), with high correlation (Pearson’s r > 0.99) and negligible mean differences (< 0.2%). Cross-platform comparison between the 3T Siemens and the 1.5T GE scanners showed mean differences of < 5%. After linear calibration, these variations were reduced to insignificant levels, yielding a strong correlation (Pearson’s r > 0.97) and mean differences within ±0.2%.

**Conclusion:** MagDensity, an MRI-based BD measure, exhibits robustness and reliability across diverse scanner models, vendors, and field strengths, marking a promising advancement towards standardizing BD measurements across multiple MRI platforms. It provides a valuable tool for monitoring subtle longitudinal changes in BD, which is vital for breast cancer prevention and personalized treatment strategies.

## Introduction

Breast cancer remains a significant global health challenge, being a leading cause of morbidity and mortality among women. Extensive research has identified various risk factors, with higher breast density (BD) recognized as one of the most crucial, closely linked to an increased risk of developing breast cancer [1,2]. BD has been incorporated into several breast cancer risk models for personalized risk assessment, such as Tyrer-Cuzick model, resulting in improved risk discrimination [3–7]. Its critical role was further highlighted by legislative reforms initiated in 2011, when more than half of the U.S. states passed laws requiring healthcare providers to inform patients with dense breasts [8]. This significant move acknowledges that higher BD increases cancer risk and may necessitate supplemental screening measures [9]. Moreover, longitudinal changes in BD have been examined in clinical trials as surrogate measures for evaluating the efficacy of hormone therapies (e.g., tamoxifen and aromatase inhibitors) used for the prevention and treatment of breast cancer [10,11]. A previous study demonstrated that a reduction in mammographic density after 12– 18 months of tamoxifen therapy was associated with a reduction in cancer risk [12]. Thus, a sensitive BD measurement is not only useful for evaluating the risk of developing breast cancer, but also for monitoring the effectiveness of preventive interventions and tailoring treatment strategies for individual patients.

The current standard of care for assessing BD includes mammography, followed by a qualitative categorization into one of the four major categories according to the Breast Imaging Reporting and Data System (BIRADS): almost entirely fatty, scattered areas of fibroglandular tissue, heterogeneously dense breast, and extremely dense breasts. However, its inherent subjectivity introduces potential variability in interpretation [13,14]. Additionally, the reliability of BD measurements using mammography is compromised by two-dimensional projections and inconsistent breast compression; even with the support of digital detector/image enhancement algorithms or tomosynthesis, it remains challenging for mammography to accurately reflect the status of dense breast tissue [15] and is insensitive to minor BD changes. Moreover, the discomfort of breast compression and the ionizing radiation associated with mammogram examinations can negatively impact clinical trial recruitment.

Breast magnetic resonance imaging (MRI) has emerged as a promising contender for these challenges due to its distinct advantages including three-dimensional (3D) capability that avoids breast compression, strong visual contrast between fibroglandular and fatty tissues, and the absence of exposure to ionizing radiation. A previous study proposed a quantitative MRI-based BD measure known as MagDensity [16], which utilizes the Dixon fat-water decomposition method and a whole breast segmentation strategy [17]. This technique has demonstrated its reliability and quantitative accuracy in BD measurement, showing reliable results when compared to mammographic density. It has been successfully used as an outcome measure in several clinical trials (clinical trial numbers: NCT01761877 and NCT02028221) [18,19] to assess longitudinal changes in BD.

Reproducibility is a fundamental aspect of scientific research, emphasized by the National Institutes of Health (NIH) as a means to ensure that biomedical research findings can be reliably utilized in clinical settings. For women, particularly those at an elevated risk of breast cancer, who may need multiple MRI scans at different healthcare facilities over their lifetime, the consistency of BD measurements across different scanner models, field strengths, and vendors becomes paramount for clinical application [20]. The generalizability of BD measurements across diverse MRI systems is critical for their integration into standard clinical workflows, which facilitates consistent breast cancer risk assessment and monitoring across varied medical environments. A cross-scanner study is needed to ensure the robustness of BD measurements across different platforms. However, scheduling scans for different scanners on the same day poses significant challenges due to the need for precise coordination and availability of multiple MRI systems within a narrow timeframe. This requirement is crucial for a successful cross-scanner study because factors such as menstrual cycle phases and weight fluctuations can affect BD measurements, thereby undermining the reliability of the analysis [21].

In this study, we aimed to evaluate the reliability of MagDensity for BD quantification using data acquired within a three-hour window (for each participant) from three scanners with different field strengths (3T and 1.5T) and vendors (Siemens and GE). To our knowledge, this work is among the first to conduct a cross-scanner reliability assessment for a quantitative MRI-based BD measurement.

## Materials and Methods

### Study Population

This prospective study was approved by the local Institutional Review Board, and written informed consent was obtained from all participants. The recruitment period began on August 1, 2019 and concluded on November 8, 2019. All assessments and analyses were performed in accordance with relevant guidelines and regulations. Inclusion criteria of the study were: i) females within the age range of 18 to 75 years; ii) no prior history of breast cancer or known breast-related diseases, iii) no contraindications to MRI, and iv) ability to provide informed consent. Exclusion criteria included: i) long-term use of non-steroidal anti-inflammatory drugs (NSAIDs), ii) claustrophobia or an elevated risk of cardiac arrest, iii) unable to lie comfortably on scanner bed for 20 minutes, iv) contraindication to MRI, and iv) cognitively impaired or not able to provide informed consent.

### MRI Protocol

Each participant underwent three fat-water breast MRI scans using 3T scanners (Siemens Prisma and Siemens Biograph mMR) and a 1.5T scanner (GE Signa). All participants were scanned in the prone position, identical to current practice for breast MRI. Data for each participant were acquired within a three-hour window to mitigate the impact of physiological variations, such as menstrual cycle fluctuations and weight changes. The scanning parameters for each scanner are summarized in Table 1. Note that the variations in scanning parameters were intentionally introduced as part of the study’s methodology to assess the reliability of our BD assessment technique. The sequences and parameters were chosen based on existing clinical sequences.

**Table 1.** Scanning parameters used for acquisition.

| Scanner<br>Parameter | 3T Siemens Prisma | 3T Siemens Biograph mMR | 1.5T GE Signa HDxt |
| --- | --- | --- | --- |
| **Coil** | 16-channel breast (Sentinelle, Erlangen, Germany) | 8-channel breast (Sentinelle, Erlangen, Germany) | Sentinelle 4-channel biopsy table (Sentinelle, Erlangen, Germany) |
| **Orientation** | Axial | Axial | Axial |
| **Sequence** | 3D Cartesian six-echo GRE | 3D Cartesian six-echo GRE | EFGRE3D six-echo |
| **Acquisition matrix** | $256 \times 152$ | $256 \times 152$ | $512 \times 372$ |
| **Pixel size** | $1.97 \times 1.97 \text{ mm}^2$ | $1.97 \times 1.97 \text{ mm}^2$ | $0.625 \times 0.625 \text{ mm}^2$ |
| **Flip angle** | 6 degrees | 6 degrees | 12 degrees |
| **Slice thickness/gapping** | 4/0 mm | 4/0 mm | 2/0 mm |
| **Phase encoding direction** | A – P | A – P | A – P |
| **Number of averages** | 1 | 1 | 1 |
| **Repetition time** | 21.0 ms | 21.0 ms | 22.4 ms |
| **Echo time** | 1.37, 2.66, 4.92, 6.15, 7.38, 8.81 ms | 1.37, 2.66, 4.92, 6.15, 7.38, 8.81 ms | 2.88, 6.04, 9.20, 12.35, 15.51, 18.66 ms |

GRE: gradient echo; EFGRE3D: enhanced fast gradient echo three-dimensional.
\* Note that the variations in scanning parameters were intentionally introduced as part of the study's methodology to assess the reliability of our BD assessment technique.

### Image Processing

The magnitude and phase images extracted from the scanner were combined to generate complex images. A fat-water separation technique called iterative decomposition of water and fat with echo asymmetric and least-squares estimation (IDEAL) [22] was used to reconstruct fat-only and water-only images from multi-echo data, as well as fat-water ratio maps (representing the relative percentage amount of fat signals in each voxel). Phase correction was applied to the linear and constant phase components. The VarPro algorithm [23] was employed for fat-water separation and image reconstruction, which distinguishes water and fat using complex signal fitting (rather than using magnitude data alone), accommodating multi-peak fat signals, R2* decay, and field map correction. The image processing steps were performed using in-house software developed in Matlab R2020b (MathWorks, Natick, MA) [16].

### Whole-breast segmentation

We employed an established automated whole-breast segmentation algorithm [17,24] to delineate breast regions. The algorithm included finding the most similar breast templates from a previously built dictionary, followed by an image registration step to register the chosen templates to the acquired breast data, while concurrently applying the transformations to the associated template masks to generate the masks. Introducing image registration in the segmentation process represents a significant advancement over previous methods, providing a more reliable foundation for subsequent analyses [17,24]. All procedures were executed in Matlab R2020b (MathWorks, Natick, MA). For an in-depth explanation, please refer to ref [17] for details.

### MRI-based BD Measure – MagDensity

MagDensity was calculated for the data obtained from the three scanners, with each individual breast (left or right) assessed separately. This novel MRI-based BD measurement takes into consideration both the volume and distribution of fibroglandular tissue within the breast. The cornerstone of this measurement, referred to as “FraWater”, quantifies the volumetric water fraction by correcting the fat-water signal bias – due to various factors including proton density difference, T1 effect, water content in fatty tissue - in the fat fraction maps.

In brief, a linear model is employed so that for each pixel in the breast:

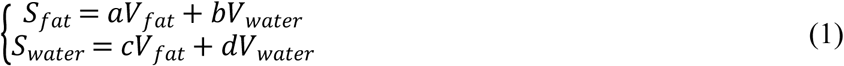

Here, S_fat_ and S_water_ are the signal intensities of fat and water, respectively; V_fat_ and V_water_ are the fat and water volume, respectively; and a, b, c, d are the correction factors. After determining the values of a, b, c, and d utilizing signal intensities from regions of “pure” fat (subcutaneous fat; V_fat_ =1, and V_water_ =0) and “pure” water (lean muscle; V_fat_ =0, and V_water_ =1) on the fat fraction map, the “FraWater”, indicating the volumetric water fraction throughout the breast, can be calculated. This modification enhances the precision of the assessment of the volume of water signal within the breast, thereby improving BD assessment. For further details, please refer to ref [16]. Combined with the registration-based breast segmentation approach, a more comprehensive and robust pipeline is offered.

### Statistical Analysis

Agreement between the MagDensity measures across the scanners was assessed using mean differences, two-tailed t-tests, Pearson’s correlation, and Bland-Altman analysis. The significance level was set at 0.05. The analyses were performed using Matlab R2020b (MathWorks, Natick, MA).

## Results

Ten healthy female volunteers aged 19–29 years (22.7 ± 3.3), with no known breast disease, were enrolled in the study. A total of 20 single breast data (left + right) were obtained. Fig 1 shows the multi-echo images from the three scanners for a representative volunteer. Fig 2 displays the reconstructed fat-only and water-only images utilizing the echo images, along with the corresponding fat fraction maps used for MagDensity calculation.

**Fig 1.**
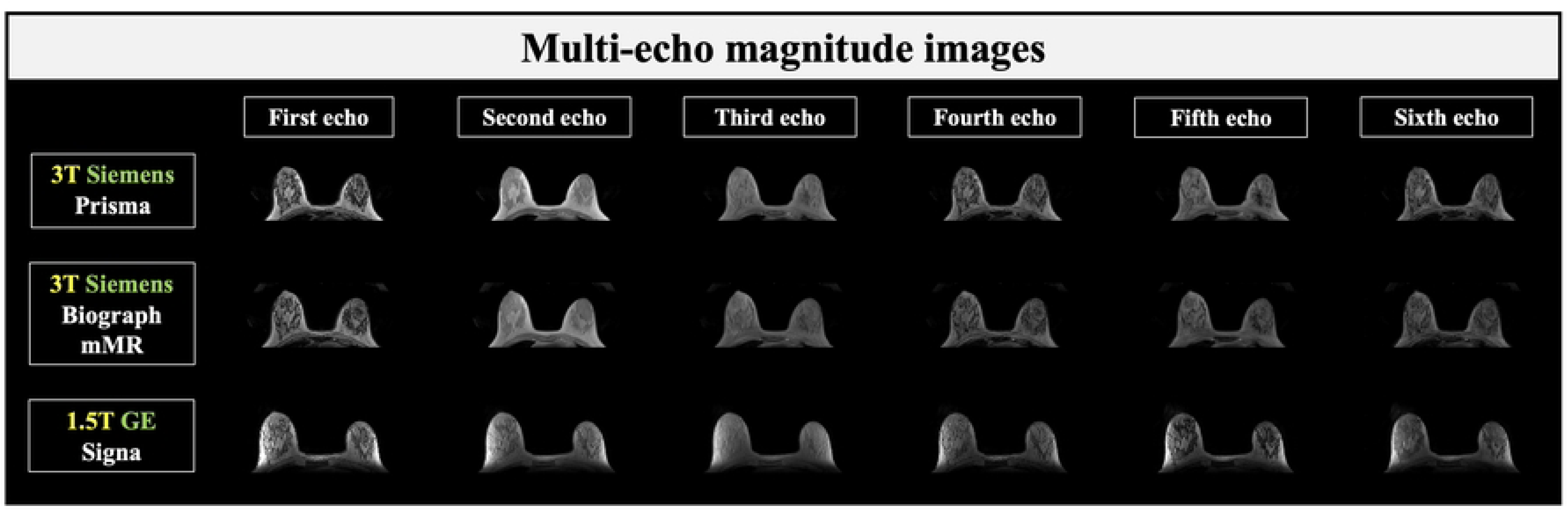
Multi-echo magnitude images from different scanners. Multi-echo magnitude images acquired from a representative participant, using 3T Siemens Prisma (top row), 3T Siemens Biograph mMR (middle row), and 1.5T GE Signa (bottom row).

**Fig 2.**
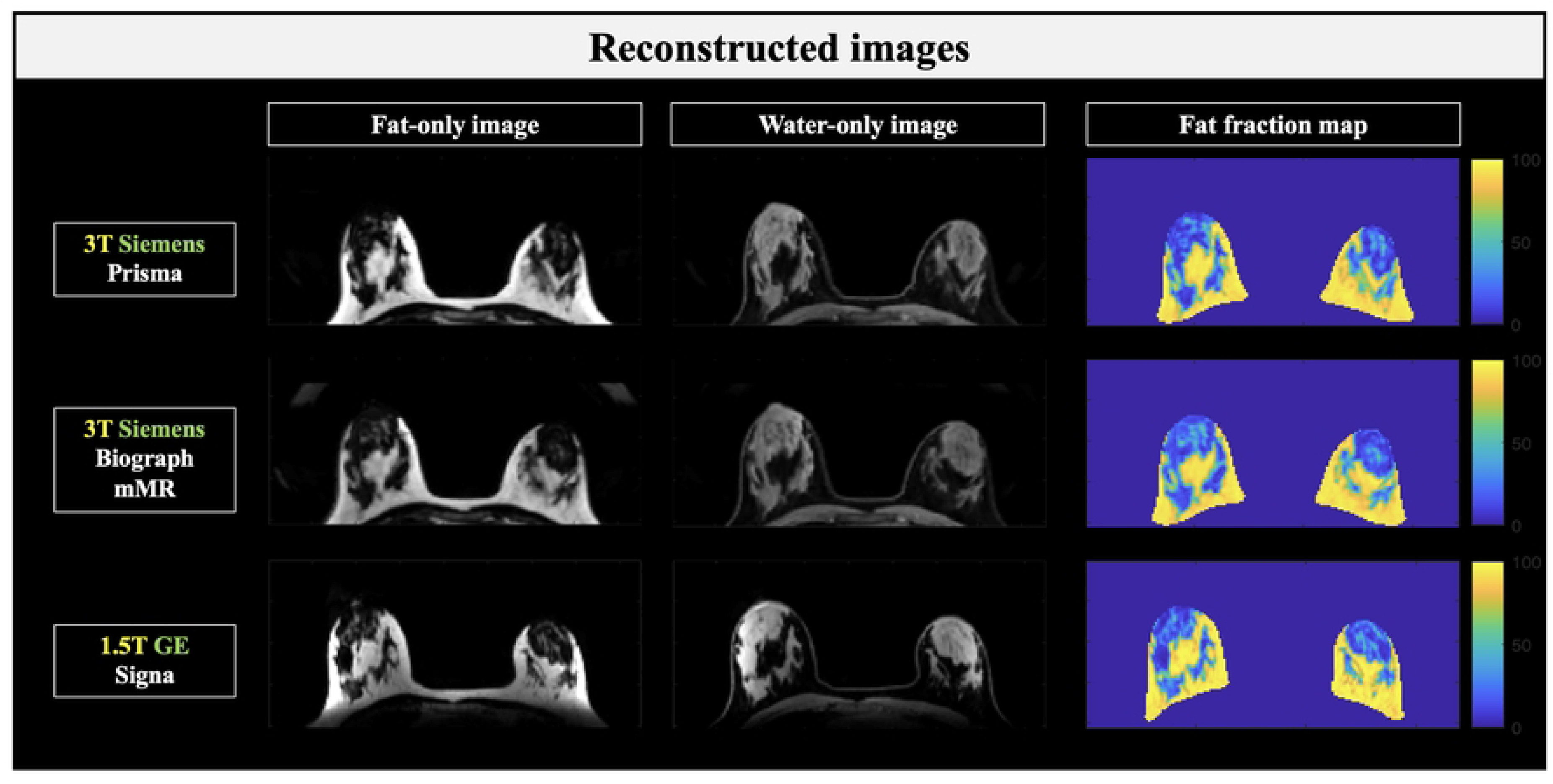
Reconstructed images using the multi-echo data. Reconstructed images from a representative participant, including fat-only images (left column), water-only images (middle column), and regions of interest on the fat fraction maps (right column) used for MagDensity calculations.

For MagDensity measures from the same field strength (3T) and the same vendor (Siemens), the observed cross-scanner mean differences were small (within 0.2%). Pairwise t-test found no statistically significant difference between the MagDensity measures of 3T Siemens Prisma and 3T Siemens Biograph mMR (*p* > 0.05). The results are summarized in Table 2. Fig 3 presents the Pearson’s correlation and Bland–Altman plots.

**Fig 3.**
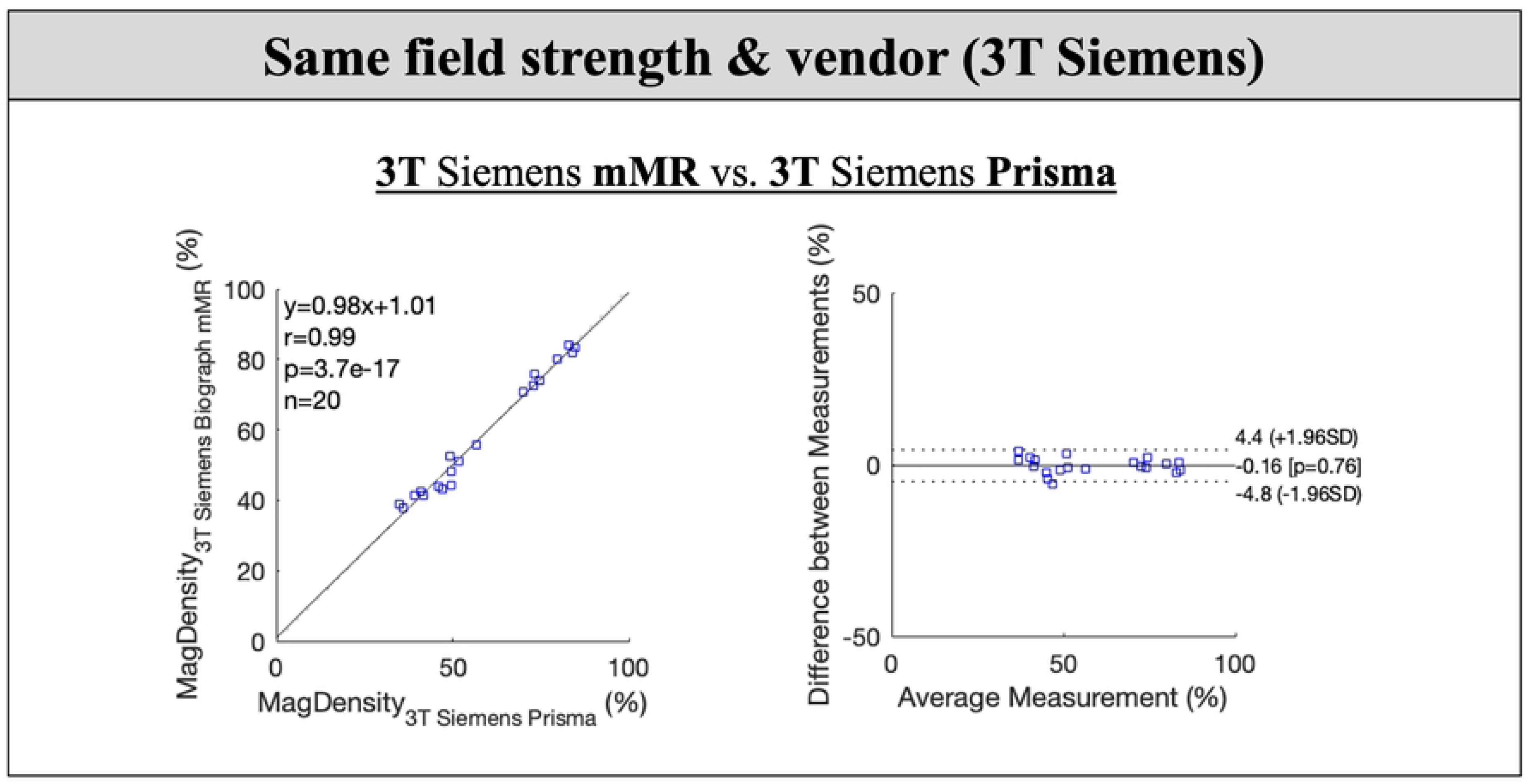
Pearson’s correlation and Bland–Altman analyses between the MagDensity measures of 3T Siemens Prisma and 3T Siemens Biograph mMR. Pairwise t-test showed no statistically significant difference between the MagDensity measures (p > 0.05) with a Pearson coefficient larger than 0.99 (p < 0.001). The Bland-Altman analysis showed a mean bias of −0.16% with 95% limits of agreement between −4.8% and 4.4%.

**Table 2.**
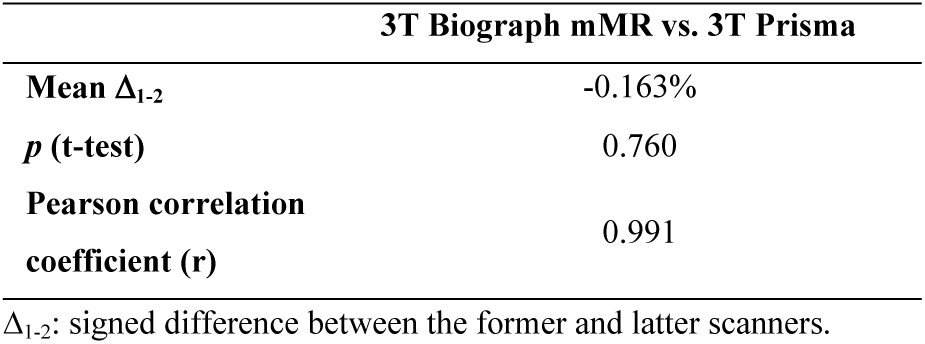
Comparison of MagDensity measures between the same field strength (3T) and the same vendor (Siemens)

For MagDensity measures across different field strengths and vendors (3T Siemens vs. 1.5T GE) as well as with different types of sequences, although they remained highly correlated (r >0.97), slightly larger mean differences were observed. However, this bias can be easily corrected by a linear calibration between the MagDensity measures of 3T Siemens Biograph mMR vs. 1.5T GE Signa (no statistically significant difference between the two Siemens scanners). All calibrated 1.5T GE Signa MagDensity values were obtained using leave-one-out cross-validation. After the calibration, the observed cross-field/vendor mean differences were within ±0.2%. Pairwise analysis showed no statistically significant difference between the MagDensity measures between 3T Siemens Prisma/Biograph mMR and 1.5T GE Signa (*p* > 0.05). The results are summarized in Table 3. Fig 4 shows the Pearson’s correlation and Bland–Altman analyses.

**Fig 4.**
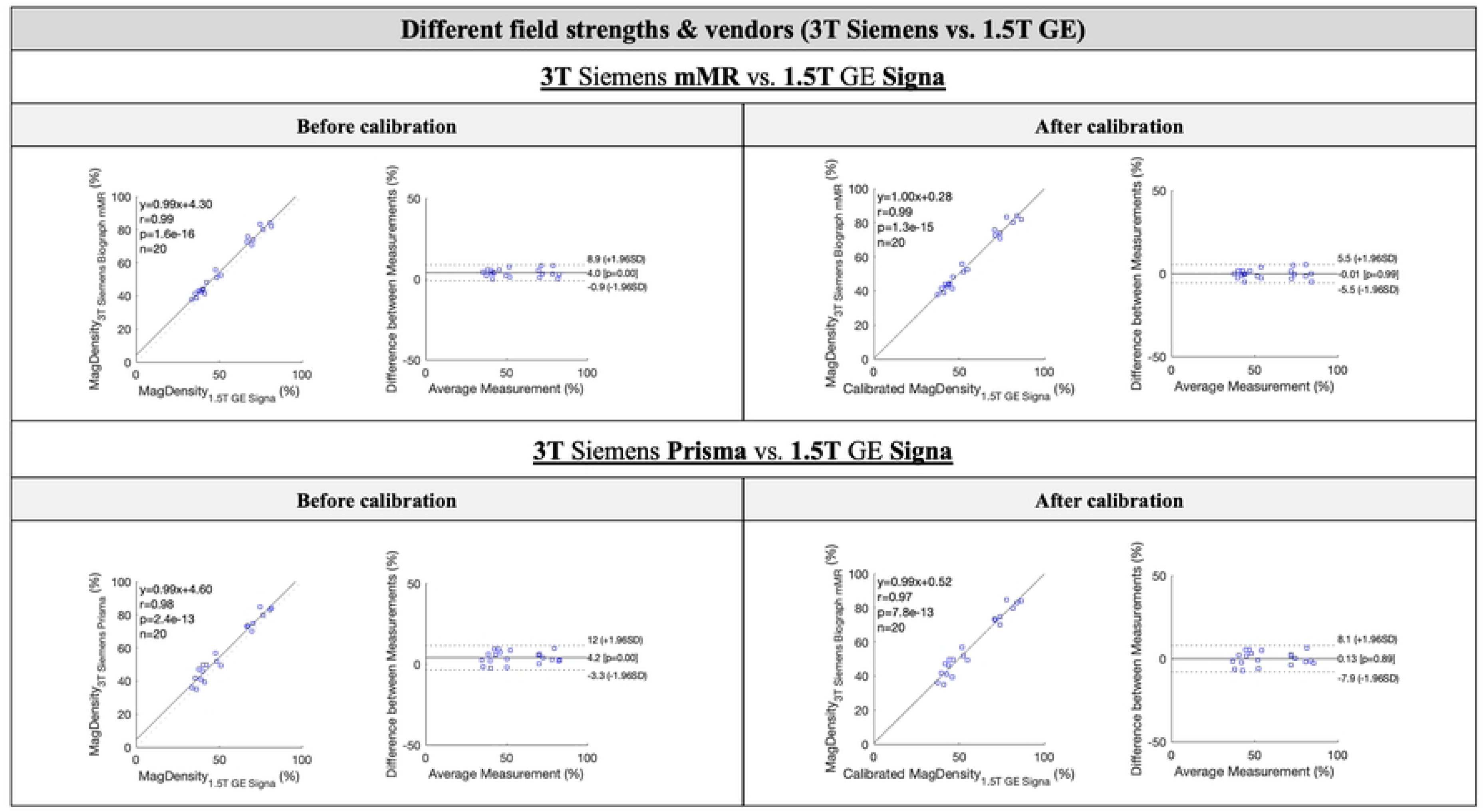
Pearson’s correlation and Bland-Altman analyses between the MagDensity measures of 3T Siemens Biograph mMR/Prisma vs. 1.5T GE Signa (before and after calibration). After calibration, pairwise t-test showed no statistically significant difference between the MagDensity measures between 3T Siemens Biograph mMR/Prisma and 1.5T GE Signa (p > 0.05) with Pearson coefficients larger than 0.97 (p < 0.001). The calibration was performed in a leave-one-out manner. For measures between 3T Siemens Biograph mMR and 1.5T GE Signa, the Bland-Altman analysis showed a mean bias of −0.01% with 95% limits of agreement between −5.5% and 5.5%. For measures between 3T Siemens Prisma and 1.5T GE Signa, the Bland-Altman analysis showed a mean bias of 0.13% with 95% limits of agreement between −7.9% and 8.1%.

**Table 3.** Comparison of MagDensity measures across different field strengths and vendors (3T Siemens vs. 1.5T GE) before and after calibration.

|  | 3T Biograph mMR vs. 1.5T Signa |  | 3T Prisma vs. 1.5T Signa |  |
| --- | --- | --- | --- | --- |
|  | Before calibration | After calibration | Before calibration | After calibration |
| Mean $\Delta_{1-2}$ | 4.023% | -0.007% | 4.186% | 0.129% |
| $p$ (t-test) | <0.001 | 0.991 | <0.001 | 0.889 |
| Pearson correlation coefficient (r) | 0.989 | 0.986 | 0.976 | 0.972 |

$\Delta_{1-2}$ : signed difference between the former and latter scanners.
\* Note that the calibration was performed in a leave-one-out manner.

## Discussion

Breast density (BD) has gained prominence as a crucial factor in the risk determination for breast cancer. In this study, we investigated the validity and reliability of the quantitative MRI-based BD measure, MagDensity, across three different scanners across two different vendors (Siemens vs. GE) and field strengths (3T vs. 1.5T), highlighting its potential for broader clinical adoption.

Our results revealed that when comparing MagDensity measures between scanners of the same vendor, sequence, and field strength (i.e., 3T Siemens), negligible differences were observed. The strong correlation (r = 0.99, *p* < 0.001) suggests consistent performance within the same technological environment and high intra-vendor reliability, affirming findings from previous test-retest studies [16]. Expanding the analysis to include different field strengths, vendors, and sequences (i.e., 3T Siemens vs. 1.5T GE) unveiled a small bias, suggesting small initial inter-vendor, -field strength, and -sequence variability. This serves as the “worst-case” scenario as the images used for MagDensity calculation were acquired on different scanners, at different field strengths, and with different acquisition sequences and parameters. This bias can be effectively addressed through linear calibration between the measures of the 3T Siemens Biograph mMR and the 1.5T GE Signa, leading to aligned values within a margin of ±0.2%. It’s essential to emphasize that our primary objective is not to assert the absolute uniformity or direct equivalence of BD measurements across diverse scanners and configurations. Rather, our primary aim is to illustrate the potential for addressing the observed variations. In essence, while we acknowledge the inherent intricacies arising from alterations in field strength, scanning protocols, and scanner vendors, our study underscores the feasibility of mitigating these complexities through a calibration approach. This methodology ensures that BD measurements obtained across different platforms maintain a consistent level of reliability, even in scenarios where multiple variables undergo simultaneous adjustments during the scanning process.

Another potential approach for calibration could be to develop a calibration phantom. By scanning the phantom on different scanners, a calibration algorithm could be built. While this approach offers potential, it presents significant logistical and practical challenges, such as the need for specialized fabrication, rigorous validation to ensure long-term accuracy and stability, and ongoing maintenance. Considering these substantial requirements and the early-stage nature of MagDensity’s implementation, this strategy was deemed outside the scope of our current study. Nonetheless, as MRI-derived breast density assessment advances towards broader clinical adoption, incorporating calibration phantoms into routine workflows could become a valuable future enhancement for further refining cross-platform consistency.

The exploration of BD as a predictive marker for breast cancer risk has seen considerable attention in the scientific community over the past years. Mammographic density assessment, following the BI-RADS categorization, has been the mainstay of clinical practice. Studies by Portnow et. al. [25] and Harkness [26] emphasized the inherent subjectivity associated with such qualitative categorizations, which can lead to potential variabilities in clinical interpretation. In fact, a multicenter study by Sprague et. al. [13] reported discrepancies in mammographic density categorization among radiologists, further underscoring the need for a more objective measure. Our research on the MRI-based MagDensity measure builds on these findings, proposing a more quantitative alternative that reduces human interpretive errors. In comparison to other MRI-based methodologies, our approach using the Dixon fat-water decomposition technique and whole breast segmentation offers enhanced precision as the Dixon method based on chemical shift imaging has shown to be effective for fat suppression in tissues with similar amounts of lipid and water (e.g., fibroglandular tissue) and insensitive to B0 inhomogeneities [15,22,27,28]. While Wengert et. al [28] developed an automated MRI-based BD assessment using the Dixon technique, our study distinguishes itself by not only correcting the inherent bias when calculating fat/water fractions, but also by testing and validating the technique across different scanner models, vendors, and field strengths. This cross-platform validation is crucial for ensuring the reliability and accuracy of BD measurements over time. It is particularly important in monitoring high-risk women or evaluating the impact of interventions, especially in postmenopausal women where changes are expected to be smaller in response to endocrine therapies [18]. Additionally, validated cross-platform measurements enable pooling data from different studies or centers, increasing the statistical power and generalizability of findings related to BD changes. Last but not least, BD can be influenced by various hormonal and physiological factors, including menopause, pregnancy, and lactation. Longitudinal tracking can provide insights into the effects of these life stages on breast density, thereby contributing to a more comprehensive understanding of individual breast health dynamics.

Nevertheless, the study has its limitations. One major limitation is the small sample size, consisting of only ten healthy volunteers. While this may seem insufficient for a comprehensive analysis, it is important to recognize the significant logistical challenges inherent in conducting a cross-scanner study. Coordinating scans on the same day across different MRI systems requires precise scheduling and availability, which is difficult to achieve. Despite the modest sample size, the insights gained from this initial cohort are still valuable, providing important preliminary data on the consistency of MagDensity measurements across different scanners. Future studies with larger and more diverse populations are warranted. Additionally, we have retrospectively identified that the subjects recruited for this study were of high BD (all greater than 40%). This was likely because the sample of healthy volunteers consisted mostly of young medical professionals, as the recruitment flyer was posted in the local medical school and hospital. Future studies will aim to recruit subjects with lower expected BD (e.g., post-menopause, higher weight).

Furthermore, while we used three popular scanner variants, other models and vendors might exhibit different biases that would require further exploration. Moreover, although the current standard of care still relies on the BIRADS categories, there does exist automated software for quantitative mammographic BD assessment (e.g., Quantra [29], Volpara [30]); future studies could include a comparison of automated mammographic density measurements to MagDensity.

## Conclusion

In conclusion, the quantitative measure of MRI-based BD (MagDensity) exhibited high robustness within the same field strength and vendor (different models), and it demonstrated promising reliability after leave-one-out calibration across different vendors, scanner models, and field strengths. This technique offers an encouraging step towards standardizing BD measurements across various MRI platforms, which could facilitate broader clinical adoption. Future studies might focus on extending this analysis to a larger cohort across a broader array of MRI platforms.

## Data Availability

The data presented in this study will be publicly available on our lab’s website under the "Research/Available Data" section: https://sites.google.com/stonybrook.edu/miralab/research/available-data

## Acknowledgements

None.

